# Timing of non-pharmaceutical interventions to mitigate COVID-19 transmission and their effects on mobility: A cross-country analysis

**DOI:** 10.1101/2020.05.09.20096420

**Authors:** Amit Summan, Arindam Nandi

## Abstract

Non-pharmaceutical interventions (NPIs) that encourage physical distancing can decrease and delay the transmission of COVID-19. They have been implemented globally during the pandemic, however, the specific NPIs implemented and the timing of interventions has varied widely. We validated two published datasets on the implementation of NPIs globally. The health and socioeconomic factors associated with delay in implementation of NPIs was analyzed using fractional logit and probit models, and beta regression models. The probability of timely NPI implementation by a country was analyzed using a probit model. The effects of these interventions on mobility changes using Google social mobility reports, were analyzed with propensity score matching methods. Three NPIs were analyzed: national school closure, national lockdown, and global travel ban. Countries with higher incomes, larger populations, and better health preparedness measures had greater delays in implementation. Countries with greater population density, more democratic political systems, lower case detection capacity, and later arrival of first cases were more likely to implement NPIs. Implementation of lockdowns significantly reduced physical mobility. Mobility was further reduced when lockdowns were enforced with curfews or fines, or were more strictly defined. National school closures did not significantly change mobility. The implementation of NPIs is a global public good during pandemics, and the international community needs to address constraints and design incentives so countries implement NPIs in a timely manner. Further analysis is needed on the effect of NPI variations on mobility and transmission, and their associated costs.

## INTRODUCTION

As of May 6th, 2020, the novel coronavirus (SARS-COV-2) had spread to 212 countries and territories, infected over 3.75 million people, and caused over 263,000 confirmed deaths worldwide. The estimated global cost of COVID-19 without containment measures — population level social distancing along with surveillance and quarantine — is $9 trillion^1^ and the estimated death toll is 40 million^2^. Mortality would be disproportionately concentrated in those who are at greater for transmission such as frontline healthcare workers, those who have underlying risk factors such as economically disadvantaged groups and the elderly, and those with lack of access to critical care.^3,4^

National governments have taken two main approaches to limiting transmission. Countries such as South Korea, Singapore, and Germany have used intensive testing, innovative technologies to contract trace, and quarantine and isolation measures to keep cases low, along with moderate social distancing measures.^56^ However, the success of such approaches depends on early implementation. This strategy also requires robust logistics and testing capacity, which many countries may lack.^8^ Given a basic reproduction number of 2.5 and a low rate of pre-symptomatic transmission, an isolation and contact tracing approach would require tracing of an estimated 70% of contracts.^9^ A recent study showed that peak infectiousness time for COVID-19 occurs before or at symptom onset,^10^ suggesting this method alone would not suffice, or would require considerable testing capacity.

The alternative approach is the implementation of non-pharmaceutical interventions (NPIs) that encourage social distancing. NPIs in combination with widespread testing, case detection, contact tracing, and enforcement of quarantine are appropriate where there is widespread community transmission.^5,11^ These measures have delayed transmission and flattened the COVID-19 epidemiological curve, buying governments precious time to prepare for higher caseloads.^12-14^ NPIs work best when they are applied as a basket of measures — a rapid review found that quarantine combined with multiple preventive measures such as school closures, travel restrictions, and social distancing, had a larger cumulative effect on new cases, transmission rates, and number of deaths, than any single intervention alone.^15^

The effectiveness of NPIs may be a function of when they are implemented, with earlier implementation being more successful in reducing transmission.^16^ American cities that implemented multiple NPIs earlier had lower death rates during the 1918 influenza pandmeic.^17^ Countries may choose to gradually implement measures or delay implementation altogether to minimize the economic and social costs of lockdowns^18,19^, or even political costs^8^. These costs include reduced economic growth,^20^ increased risk of depression and mental health problems due to isolation,^21–24^ and increased risk of domestic violence ^25-27^. These costs may vary by country and within countries; lockdown measures can have disproportionate costs where governments are not able to provide social safety nets.^28^ A lack of knowledge about fundamental disease characteristics can also delay the most appropriate response.^29^

It is important to understand what considerations are made in the implementation and the timing of NPIs to see how resources can be better allocated and incentives more effectively created to improve NPI implementation for COVID-19 and future pandemics. Although many predictive mathematical models ^13,15,30-32^ have simulated the effect of NPIs on COVID-19, evidence is just recently becoming available on the actual effect of these interventions and the decision models that influenced their implementation. We examined country-level health systems capacity, epidemiological, and socioeconomic characteristics associated with delay in the implementation of three NPIs: national school closure, global travel ban, and country-wide lockdowns, and the effect of these NPIs on population mobility.

## DATA AND METHOD

### Data

Daily data on the number of COVID-19 cases were collected from the European Center of Disease Prevention and Control (ECDPC).^33^ Our data on NPIs were drawn from two datasets – the ACAPS COVID-19 Government Measures dataset^34^ (May 1, 2020 release) and a dataset constructed by the University of Oxford^35^ (April 29, 2020 release). Both of these datasets contain information on global, country-level COVID-19 related policy interventions and implementation dates. We carefully scrutinized the data and its sources for consistency, and checked for consistency across the two datasets. We considered three interventions: national school closure, national lockdown, and global travel ban. We did not include countries where measures were implemented only at the sub-national level (e.g. province or city).

We used Google’s recent “mobility” reports which track mobile device location data for over 130 countries as measures of social mobility and physical distancing.^36^ These reports have been available since mid-February of 2020 and have been updated on a weekly basis to aid policy makers. The reports show trends in how visits and length of stay at different places change compared to a baseline. The baseline is the median value, for the corresponding day of the week, during the period of January 3, 2020 to February 6, 2020. Google identifies six location types for which mobility data is tracked: retail and recreation, grocery and pharmacy, parks, transit stations, workplaces, and residential.

Our control variables included health systems, epidemiological, and socioeconomic characteristics which can affect disease transmission rates, epidemic costs, and cost and benefits of NPI implementation. The following variables were collected from the World Bank database^37^: log of population density, percentage of the population under the age 15, and the log of total population. We also included 8 sub-regions of the world to capture cultural or geographical factors that may affect the response to COVID-19 and country income category (low, lower middle, upper middle, and high) from the World Bank. Additionally, we included a measure of government regime type from the Center for Systemic Peace.^38^ This variable varied from −10, indicating an autocracy, to 10, indicating a full democracy. We also included the number of cases per 100,000 people two weeks after the arrival of the first case and the day the first case was detected in the country as additional control variables. Finally, as a measure of health systems capacity related to pandemics, we used the global health security index score — developed by the Nuclear Threat Initiative and the Johns Hopkins Center for Health Security — which uses 140 variables related to six categories: prevention, detection and reporting, rapid response, health system, compliance with international norms, and risk environment, to develop a country score between 0 and 100, where a higher score indicates a greater level of pandemic preparedness.^39^

### Methods

#### Outcomes

We examined three NPIs: lockdown, global travel ban (border closure to non-essential travel), and school closure. All three measures curb social interaction among individuals. Although the evidence on the effect of travel bans^40-43^ and school closures^44-47^ on transmission delay and spread is mixed, we included both of these NPIs because they had widespread implementation globally. We focused on national-level interventions as sub-national data may not always be available or complete.

We defined lockdown as the closure of all non-essential businesses and allowance of leaving home only for ‘essential’ activities’. The definition of essential activities and businesses may vary by country. For example, some countries closed all retail stores, recreational business and areas, and workplaces that may be at high risk for transmission, and recommended all others to work from home, and only go to work if *absolutely* necessary. What is considered absolutely necessary may vary by country, by employer, or to an individual. Some countries allowed for exercise outside of the home or leaving the home to get ‘fresh air’ for a limited time. To distinguish between the intensity of the lockdown, we created a measure of strict lockdown. A strict lockdown was considered one in which all industries were explicitly completely closed except for those deemed essential, that is, related to health, food, or necessary for the provision of essential goods and services (e.g. pharmacies, grocery stores, financial institutions, healthcare, food production, etc.), and individuals were only allowed to leave if they worked in one of these essential businesses without exception, or for food and medical needs including accessing care or providing care to family members.

We also considered lockdown to be strict if a normal lockdown was accompanied by one of the following: 1) a curfew which allowed individuals to engage in sanctioned activities outside the home at specific time intervals, 2) a fine which would be issued if individuals were not complying with lockdown measures, and 3) additional military presence to enforce lockdown measures. We will refer to a non-strict lockdown as a normal lockdown from hereon. We only considered lockdowns that had a minimum length of 72 hours, which excluded some countries that, for example, implemented measures during weekends or extended weekends only. We used the earliest type of lockdown implemented for a country, resulting in one observation per country. This is important because many countries started with a normal lockdown and slowly transitioned into a strict lockdown or vice-versa.

For our analysis we created three binary variables that considered if a lockdown, strict lockdown, or national school closure was implemented within a certain time frame. We also created a continuous variable that looked at the delay in implementation of either a lockdown, school closure, or global travel ban after first case detection. The outcomes variables are described in Table 1. The binary variables of NPI implementation only considered countries who had implemented the policy within the specified time period or those that had a minimum time since first case detection at least as long as the latest day a country could have implemented the NPI under consideration.

**Table 1:** Summary of outcomes variables

| Variable | Policy | Description | Maximum delay (days) in implementation |
| --- | --- | --- | --- |
| *school30* | National school closure | Coded 1 if policy implemented within specified time period. Coded 0 if days since first case were at least the length of the maximum specified time period, and the NPI was not implemented. | 30 |
| *school45* |  |  | 45 |
| *lockdown30* | National lockdown (closure of all non-essential businesses and public areas; residents strongly recommended to stay at home and leave when absolutely necessary). |  | 30 |
| *lockdown45* |  |  | 45 |
| *lockdown_strict30* | National lockdown (strict:): 1) only essential business open, not allowed to leave home except for essential needs (food, work in essential business, healthcare related needs), 2) curfew (limited times) for leaving home to perform essential activities, 3) fines if lockdown policies or violated, or 4) military presence to enforce lockdown. |  | 30 |
| *lockdown_strict45* |  |  | 45 |
| *delay1_30* | Lockdown, national school closure, or global travel ban (on non-essential travel) | Delay in days in implementation of one of the policies within the specified time period since 1st case detection. | 30 |
| *delay1_45* |  |  | 45 |

Note: For European Union countries, we considered countries that implemented a ban to at least all non-Schengen zone countries to have implemented a global travel ban.

The final set of variables measured the change in mobility. The main outcome variable was the percentage change in mobility from one day before to two days after the implementation of an NPI for each of the six locations for which Google reports data. We focused on a short post-intervention follow up period for our main analysis because the probability that a country changes recently implemented measures or implements new measures after an NPI is introduced increases with time and may make it difficult to isolate the effects of the intervention of interest. However for additional sensitivity analysis, we looked at the change in mobility from one day before to six days after NPI implementation.

#### Estimating associations with delay in policy implementation

To estimate the association of country characteristics with delay in implementation of a policy after first case detection, we employed three methodologies. The number of days in delay in implementation contains discrete values and is bounded below by 0 and above by the maximum number of days in implementation we are considering as described in Table 1. Because our outcome was bounded by a maximum number of days, we standardized our outcome variable to be between the interval [0, 1], by dividing the outcome variable by 30 or 45 days depending on the latest implementation day for countries to be considered for analysis. Then we employed fractional logit regression, and two additional models for sensitivity analysis – the fractional probit model and beta regression model. The beta distribution requires values to be bounded between (0,1) and cannot include boundary values, therefore we added epsilon (1^-10^) to transformed delay values of 0 and subtracted epsilon from transformed delay values of 1. This model was regressed on the type of NPI, region, income level, health preparedness score, log of population density, log of population, measure of government regime type, percentage of young population, and number of cases per 100,000 people two weeks after first case detection. Standard errors were clustered at the country level.

#### Propensity score matching

Propensity score matching (PSM) is a widely used statistical approach used to analyze the effects of interventions in non-experimental data. In observational settings, assignment to intervention or control groups, or self-selection into a group is not random. Assignment is typically correlated with a number of variables and a simple comparison of outcomes between intervention and control groups may produce biased estimates. PSM homogenizes the two groups by matching each intervention observation with one or more similar control observations. The difference in outcome between the two matched groups is known as the average treatment effect on the treated (ATT).

We employed a probit model to regress the binary indicator of whether a country implemented NPI on a set of covariates which included the region, income level, health preparedness score, log of population density, log of population, measure of government regime type, percentage of young population, the date of arrival of the first case, and number of cases per 100,000 people two weeks after first case detection. Based on the predicted probability (propensity score) from this regression, we matched intervention countries with untreated countries.

We used one-to-one nearest-neighbor matching with replacement. For sensitivity analyses, we matched observations to the nearest three neighbors and also employed the kernel matching algorithm. In all models, common support was imposed, where observations below the minimum or above the maximum propensity score for the treated group were dropped.

We investigated the quality of matching between the groups in three ways. First, we looked at difference in mean and median percentage bias across all matching variables before and after matching. A reduction in bias indicated the matching procedure had made the control and intervention groups more comparable. Second, we looked at the p-value of the likelihood ratio test of joint significance of all matching variables on the propensity score. Lastly, we looked at the pseudo R^2^ of this model. A higher p-value after matching or lower pseudo R^2^ would mean there was a reduction in systematic differences in variables. All analysis was conducted using Stata version 14.2 and we considered p<0.05 for statistical significance.

## RESULTS

### Summary characteristics of data

Table 2 shows the factors associated with delay in implementation of NPIs for countries after detecting their 1^st^, 5^th^, and 10^th^ COVID-19 case. This table does not consider countries that did not implement measures. We see that the mean delay for implementation of nationwide school closure is shortest among the three interventions — an average of 13 days after 1^st^ case detection. This is followed by international air travel restrictions at 18 days, and national lockdowns at 21 days after the 1^st^ case. The mean delay to implementation of any measure on average is highest for South Asia and the East Asia and Pacific regions, while it is lowest for the Sub-Saharan Africa, and Latin America and Caribbean regions. In this table we divided the continuous variables into categorical variables below and above their mean values for ease of interpretation. Countries with lower income; higher per capita cases two weeks after their first case; less democratic systems; and younger, smaller, and less denser populations implemented measures earlier on average. The difference in delay after 1^st^ case detection was greater for countries with higher health preparedness levels, 40 days for countries above and 12 days for countries below the median score; higher log of total population, 38 days for countries above and 16 days for countries below the median log population; and lower per capita cases two weeks after first case detection, 36 days for countries above and 17 days for countries below the median per capita case values.

**Table 2:** Mean delay (days) in implementation of NPI by country characteristics

|  | Delay1 | Observations | Delay5 | Observations | Delay10 | Observations |
| --- | --- | --- | --- | --- | --- | --- |
| **Intervention Type** |  |  |  |  |  |  |
| Air | 19.15 | 109 | 12.06 | 105 | 10.16 | 98 |
| Lockdown | 16.62 | 79 | 11.54 | 78 | 9.14 | 73 |
| School | 18.07 | 137 | 9.81 | 135 | 7.58 | 129 |
| **Region** |  |  |  |  |  |  |
| East Asia & Pacific | 11.92 | 30 | 9.85 | 27 | 8.69 | 27 |
| Europe & Central Asia | 19.35 | 100 | 12.32 | 100 | 10.26 | 100 |
| Latin America & Caribbean | 5.43 | 60 | 3.56 | 60 | 2.43 | 58 |
| Middle East & North Africa | 6.29 | 41 | 3.94 | 41 | 3.01 | 41 |
| North America | 0.68 | 3 | 0.54 | 3 | 0.31 | 3 |
| South Asia | 4.51 | 12 | 1.07 | 12 | 0.79 | 10 |
| Sub-Saharan Africa | 5.66 | 79 | 2.13 | 75 | 1.39 | 61 |
| **Income Level** |  |  |  |  |  |  |
| Low Income | 3.80 | 44 | 0.78 | 40 | 0.44 | 31 |
| Lower Middle Income | 8.93 | 67 | 3.45 | 64 | 2.63 | 60 |
| Upper Middle Income | 16.51 | 102 | 11.61 | 102 | 9.58 | 97 |
| High Income | 24.60 | 112 | 17.57 | 112 | 14.23 | 112 |
| **Growth in cases per capita** |  |  |  |  |  |  |
| Below median | 36.37 | 175 | 21.06 | 171 | 16.80 | 165 |
| Above median | 17.03 | 135 | 12.23 | 135 | 10.08 | 131 |
| **Political system** |  |  |  |  |  |  |
| Below median | 22.31 | 144 | 13.24 | 137 | 10.85 | 126 |
| Above median | 28.52 | 154 | 18.34 | 154 | 14.64 | 147 |
| **Young population** |  |  |  |  |  |  |
| Below median | 35.96 | 164 | 25.59 | 164 | 21.28 | 164 |
| Above median | 17.15 | 153 | 7.51 | 146 | 5.34 | 128 |
| **Log population** |  |  |  |  |  |  |
| Below median | 16.31 | 130 | 9.86 | 129 | 7.80 | 118 |
| Above median | 37.50 | 193 | 23.55 | 187 | 19.08 | 180 |
| **Population density** |  |  |  |  |  |  |
| Below median | 21.08 | 168 | 12.27 | 165 | 9.26 | 156 |
| Above median | 32.39 | 149 | 21.08 | 147 | 17.59 | 138 |
| **Health preparedness** |  |  |  |  |  |  |
| Below median | 12.43 | 130 | 5.10 | 123 | 3.39 | 107 |
| Above median | 40.01 | 182 | 27.39 | 182 | 22.70 | 180 |

Note: *Delay1* = delay in days in implementation of one of three NPIs after detection of 1st case. If policy was implemented before the 1<sup>st</sup>, 5<sup>th</sup>, or 10<sup>th</sup> case was detected, a delay value of 0 was used.

### Delay in implementation

Table 3 shows the results from the fractional logit, fractional probit, and beta regression models for the number of days it took to implement an NPI given that the country implemented the measure within 30 or 45 days. The response variable is transformed between 0 and 1, where .1 corresponds to 3 days and 4.5 days, if the policy was implemented 30 and 45 days since first case detection, respectively. According to all models, countries implemented national school closures (odds ratio [OR]=0.513, 95% Confidence Interval [CI]: 0.379-0.696, p-value<0.01; model 1) faster than other interventions. The Latin American and Caribbean had a shorter delay in implementation of NPIs (OR=0.084, CI: 0.030-0.231, p-value<0.01; model 1) relative to North America. Higher income countries and countries with greater health preparedness scores (OR=1.026, 1.006-1.048, p-value<0.05; model 1) had larger delays in implementation of NPIs. Greater growth in cases per capita in the first two weeks since first case detection decreased implementation delay (OR=0.965, 0.906-0.985, p-value<0.01; model 1). The log of total population was associated with greater delay in implementation of NPIs in only the beta regression model for countries who implemented the policy within 30 days, and for all models when considering countries who implemented the policy within 45 days.

**Table 3:** Country characteristics and delay in NPI implementation

| Maximum delay in implementation (days) | 30 |  |  | 45 |  |  |
| --- | --- | --- | --- | --- | --- | --- |
| Model | 1 | 2 | 3 | 4 | 5 | 6 |
|  | Fractional logit | Fractional probit | Beta regression | Fractional logit | Fractional probit | Beta regression |
| *Intervention (Air=0)* |  |  |  |  |  |  |
| Lockdown | 1.145<br>(0.846 - 1.549) | 1.091<br>(0.909 - 1.311) | 1.762*<br>(1.095 - 2.835) | 1.025<br>(0.825 - 1.273) | 1.021<br>(0.893 - 1.167) | 1.365<br>(0.916 - 2.034) |
| School | 0.513**<br>(0.379 - 0.696) | 0.667**<br>(0.555 - 0.801) | 0.575*<br>(0.332 - 0.994) | 0.605**<br>(0.470 - 0.780) | 0.740**<br>(0.637 - 0.861) | 0.581*<br>(0.361 - 0.936) |
| *Region (North America=0)* |  |  |  |  |  |  |
| East Asia & Pacific | 0.315*<br>(0.099 - 0.995) | 0.552<br>(0.105 - 2.899) | 1.24<br>(0.434 - 3.547) | 0.161<br>(0.013 - 2.079) | 0.385<br>(0.090 - 1.653) | 0.968<br>(0.353 - 2.654) |
| Europe & Central Asia | 0.150**<br>(0.053 - 0.424) | 0.352<br>(0.064 - 1.920) | 0.578<br>(0.203 - 1.647) | 0.107+<br>(0.009 - 1.314) | 0.304<br>(0.072 - 1.279) | 0.676<br>(0.270 - 1.692) |
| Latin America & Caribbean | 0.084**<br>(0.030 - 0.231) | 0.247<br>(0.044 - 1.388) | 0.289**<br>(0.116 - 0.723) | 0.056*<br>(0.004 - 0.773) | 0.205*<br>(0.046 - 0.925) | 0.281**<br>(0.129 - 0.611) |
| Middle East & North Africa | 0.281*<br>(0.104 - 0.759) | 0.517<br>(0.087 - 3.085) | 1.271<br>(0.458 - 3.527) | 0.138<br>(0.010 - 1.970) | 0.354<br>(0.077 - 1.628) | 1.223<br>(0.572 - 2.612) |
| South Asia | 0.44<br>(0.440 - 0.440) | 0.686<br>(0.087 - 5.428) | 1.222<br>(0.265 - 5.624) | 0.231<br>(0.010 - 5.354) | 0.493<br>(0.081 - 3.006) | 2.121<br>(0.468 - 9.614) |
| Sub-Saharan Africa | 0.133**<br>(0.053 - 0.334) | 0.332<br>(0.054 - 2.025) |  | 0.087+<br>(0.005 - 1.385) | 0.272<br>(0.056 - 1.318) |  |
| *Income level (Low income=0)* |  |  |  |  |  |  |
| Lower middle income | 0.993<br>(0.510 - 1.933) | 0.991<br>(0.671 - 1.462) | 1.789<br>(0.729 - 4.392) | 1.015<br>(0.565 - 1.822) | 1.001<br>(0.721 - 1.389) | 1.687<br>(0.742 - 3.837) |
| Upper middle income | 2.155+ | 1.575+ | 3.023* | 1.986+ | 1.486* | 2.677* |
|  | (0.947 - 4.903) | (0.971 - 2.556) | (1.056 - 8.652) | (0.997 - 3.958) | (1.004 - 2.198) | (1.074 - 6.674) |
| High income | 2.300+ | 1.644+ | 2.527 | 2.331+ | 1.654* | 2.514+ |
|  | (0.862 - 6.134) | (0.919 - 2.940) | (0.732 - 8.721) | (0.976 - 5.571) | (1.003 - 2.730) | (0.852 - 7.422) |
| Cases per 100,00, at 2 weeks | 0.945** | 0.964** | 0.934* | 0.939** | 0.962** | 0.919** |
|  | (0.906 - 0.985) | (0.940 - 0.989) | (0.880 - 0.990) | (0.896 - 0.983) | (0.937 - 0.988) | (0.867 - 0.974) |
| Political regime | 1.026 | 1.016+ | 1.022 | 1.007 | 1.005 | 1.02 |
|  | (0.994 - 1.059) | (0.997 - 1.035) | (0.989 - 1.057) | (0.979 - 1.035) | (0.989 - 1.021) | (0.988 - 1.054) |
| % of population under 15 | 1.009 | 1.005 | 0.971 | 0.996 | 0.997 | 0.966 |
|  | (0.976 - 1.044) | (0.985 - 1.026) | (0.926 - 1.018) | (0.965 - 1.028) | (0.979 - 1.016) | (0.925 - 1.009) |
| Log of population | 1.117 | 1.067 | 1.275* | 1.198* | 1.109* | 1.273** |
|  | (0.946 - 1.320) | (0.966 - 1.179) | (1.056 - 1.539) | (1.029 - 1.395) | (1.015 - 1.212) | (1.077 - 1.506) |
| Log of population density | 1.074 | 1.043 | 1.219+ | 1.072 | 1.04 | 1.208+ |
|  | (0.919 - 1.256) | (0.949 - 1.147) | (0.994 - 1.494) | (0.923 - 1.245) | (0.953 - 1.135) | (0.990 - 1.473) |
| Health preparedness | 1.026* | 1.016* | 1.026+ | 1.020* | 1.011* | 1.027* |
|  | (1.006 - 1.048) | (1.003 - 1.029) | (0.998 - 1.055) | (1.001 - 1.039) | (1.000 - 1.022) | (1.001 - 1.053) |
| Observations | 232 | 232 | 232 | 248 | 248 | 248 |
| Pseudo R-squared | 0.1 | 0.1 |  | 0.09 | 0.09 |  |

Note: Outcome variable is the number of delays in lockdown on a scale of (0,1) where 0 corresponds to 0 day delay and 1 corresponds to the maximum allowed delay (30 or 45 days). If policy was implemented before the first case was detected, a delay value of 0 was used. Results reported in odds ratio. Standard errors clustered at country level. 95% confidence intervals in parentheses. +p<0.1, \*p<0.05, \*\*p<0.01.

### Likelihood of implementation

Table 4 shows the likelihood of implementing school closures and lockdowns before 30 and 45 days after detecting the first case. The likelihood of implementing a normal lockdown, strict lockdown, or national school closure was not significantly associated with country’s region. Upper middle-income countries were more likely than low income countries to implement any of the three NPIs within 30 days of first case detection. More democratic countries were more likely to implement a lockdown (OR=1.048, CI: 1.001-1.097, p-value<0.05; model 1), but no significant association was found between political regime and implementation of a strict lockdown. Increased number of per-capita cases at 2 weeks after first case detection was associated with a lower likelihood of implementation for all three NPIs considered (OR=0.890, CI: 0.812-0.976, p-value<0.05; model 1), while the later the first case was detected the more likely the country was to implement any NPI. Countries that were denser were more likely to implement a lockdown, including a strict lockdown (OR=1.316, CI: 1.049-1.651, p-value<0.05; model 3), while countries with larger populations were more likely to implement only a national school closure

**Table 4:** Probability of implementing school and lockdown intervention by timeliness

| Model | 1 | 2 | 3 | 4 | 5 | 6 |
| --- | --- | --- | --- | --- | --- | --- |
| Intervention | Lockdown | Lockdown | Lockdown Strict | Lockdown Strict | School | School |
| Maximum implementation delay (days) | 30 | 45 | 30 | 45 | 30 | 45 |
| *Region (North America=1)* |  |  |  |  |  |  |
| East Asia & Pacific | 0.763<br>(0.209 - 2.780) | 1.541<br>(0.406 - 5.842) | 0.579<br>(0.171 - 1.962) | 0.769<br>(0.175 - 3.375) | 0.286+<br>(0.080 - 1.017) | 0.497<br>(0.114 - 2.164) |
| Europe & Central Asia | 1.364<br>(0.402 - 4.633) | 2.519<br>(0.715 - 8.881) | 0.525<br>(0.167 - 1.645) | 0.701<br>(0.216 - 2.278) | 0.815<br>(0.168 - 3.950) | 1.186<br>(0.205 - 6.865) |
| Latin America & Caribbean | 0.808<br>(0.294 - 2.223) | 1.188<br>(0.388 - 3.642) | 0.611<br>(0.216 - 1.728) | 0.699<br>(0.230 - 2.126) | 0.431<br>(0.138 - 1.344) | 0.492<br>(0.152 - 1.596) |
| Middle East & North Africa | 2.157<br>(0.684 - 6.802) | 2.287<br>(0.720 - 7.269) | 1.36<br>(0.465 - 3.981) | 1.15<br>(0.382 - 3.467) | 8.867*<br>(1.597 - 49.236) |  |
| South Asia | 0.598<br>(0.153 - 2.335) | 0.631<br>(0.160 - 2.479) | 1.024<br>(0.265 - 3.953) | 0.772<br>(0.184 - 3.238) | 1.717<br>(0.390 - 7.549) | 1.722<br>(0.372 - 7.971) |
| *Income group (Low income =1)* |  |  |  |  |  |  |
| Lower middle income | 3.470*<br>(1.254 - 9.599) | 2.187<br>(0.753 - 6.356) | 2.319+<br>(0.854 - 6.300) | 1.868<br>(0.649 - 5.373) | 1.276<br>(0.417 - 3.899) | 0.812<br>(0.243 - 2.714) |
| Upper middle income | 6.236**<br>(1.729 - 22.498) | 3.726+<br>(0.960 - 14.461) | 6.363**<br>(1.878 - 21.566) | 4.206*<br>(1.183 - 14.955) | 4.393*<br>(1.133 - 17.040) | 1.963<br>(0.495 - 7.791) |
| High income | 8.034*<br>(1.643 - 39.282) | 4.864+<br>(0.943 - 25.094) | 3.699+<br>(0.796 - 17.189) | 2.437<br>(0.519 - 11.446) | 7.142*<br>(1.234 - 41.330) | 7.027+<br>(0.784 - 63.000) |
| Day of first case | 1.093**<br>(1.057 - 1.129) | 1.074**<br>(1.042 - 1.106) | 1.069**<br>(1.038 - 1.101) | 1.052**<br>(1.020 - 1.085) | 1.100**<br>(1.064 - 1.136) | 1.086**<br>(1.042 - 1.132) |
| Cases per 100,000, at 2 weeks | 0.890* | 0.862** | 0.906* | 0.880* | 0.966 | 0.954 |
|  | (0.812 - 0.976) | (0.785 - 0.946) | (0.820 - 1.000) | (0.796 - 0.972) | (0.919 - 1.015) | (0.899 - 1.013) |
| Political regime | 1.048* | 1.056* | 1.013 | 1.019 | 1.043 | 1.038 |
|  | (1.001 - 1.097) | (1.004 - 1.112) | (0.969 - 1.060) | (0.970 - 1.070) | (0.985 - 1.104) | (0.968 - 1.114) |
| % of population under 15 | 0.954 | 0.97 | 0.977 | 0.978 | 1.015 | 1.004 |
|  | (0.895 - 1.018) | (0.907 - 1.038) | (0.917 - 1.041) | (0.917 - 1.043) | (0.938 - 1.098) | (0.917 - 1.100) |
| Log of population | 1.205 | 1.09 | 1.003 | 0.926 | 1.334* | 1.399* |
|  | (0.962 - 1.510) | (0.873 - 1.361) | (0.822 - 1.224) | (0.753 - 1.139) | (1.017 - 1.751) | (1.007 - 1.944) |
| Log of population density | 1.389* | 1.444** | 1.316* | 1.321* | 0.936 | 0.94 |
|  | (1.080 - 1.786) | (1.124 - 1.855) | (1.049 - 1.651) | (1.067 - 1.636) | (0.722 - 1.212) | (0.716 - 1.234) |
| Health preparedness | 0.991 | 0.983 | 1.029+ | 1.019 | 1.005 | 0.97 |
|  | (0.956 - 1.027) | (0.949 - 1.019) | (0.995 - 1.065) | (0.985 - 1.055) | (0.966 - 1.046) | (0.923 - 1.020) |
| Observations | 143 | 132 | 143 | 130 | 143 | 122 |
| Pseudo R-squared | 0.306 | 0.302 | 0.207 | 0.18 | 0.426 | 0.448 |

Note: Results reported in odds ratio. Robust standard errors. 95% confidence intervals in parentheses. +p<0.1, \*p<0.05, \*\*p<0.01

### Effect on mobility

Table 5 shows the effects of the NPIs on mobility, from one day before to two days after policy implementation. Location data suggests that there was a significant reduction in time spent outside the house from the implementation of lockdowns regardless of when they were implemented. However, if a lockdown was implemented within 30 days, a strict lockdown decreased mobility to a greater degree than a normal lockdown. For example, time spent in residential areas increased by 21% (CI: 8%-34%, p-value<0.05) under a normal lockdown relative to an increase of 30% (CI: 15%-45%, p-value<0.05) in a strict lockdown. There were substantially greater reductions in time spent outside of the home in all locations for a stricter lockdown relative to a normal lockdown. The greatest difference was for time spent in parks, where there was a reduction of 36% (CI: 7%-65%, p-value<0.05) when a strict lockdown was implemented and no significant change for a normal lockdown. When we considered all countries that implemented a lockdown within 45 days, both types of lockdowns had a reduction in mobility. However, the difference in changes in mobility between a normal lockdown and a strict lockdown decreased, with a slightly greater reduction in mobility in the normal lockdown scenario. For example, time spent in residential location increased by 39% (CI: 27%-52%, p-value<0.01) for normal lockdowns relative to increases of 35% (CI: 20%-50%, p-value<0.01) for the strict lockdown scenario. However, when we considered longer follow-up periods (one day before to six days after NPI implementation), presented in Table A7 in the supplementary appendix, we see that there is a greater reduction in mobility outside of the home regardless of when the policy was implemented in the strict lockdown scenario. There was an increase in time spent at home by 219% (CI: 150%-289%, p-value<0.01) under a strict lockdown relative to a decrease of 199% (CI: 133%-264%, p-value<0.01) under a normal lockdown. The results show that there was no significant change in mobility after implementation of a national school closure. The PSM results for matching to nearest three neighbors and kernel matching results are shown in the supplementary appendix in Tables A4 to A6. The coefficients do not vary substantially, and the results are not sensitive to the matching algorithm used.

**Table 5:**
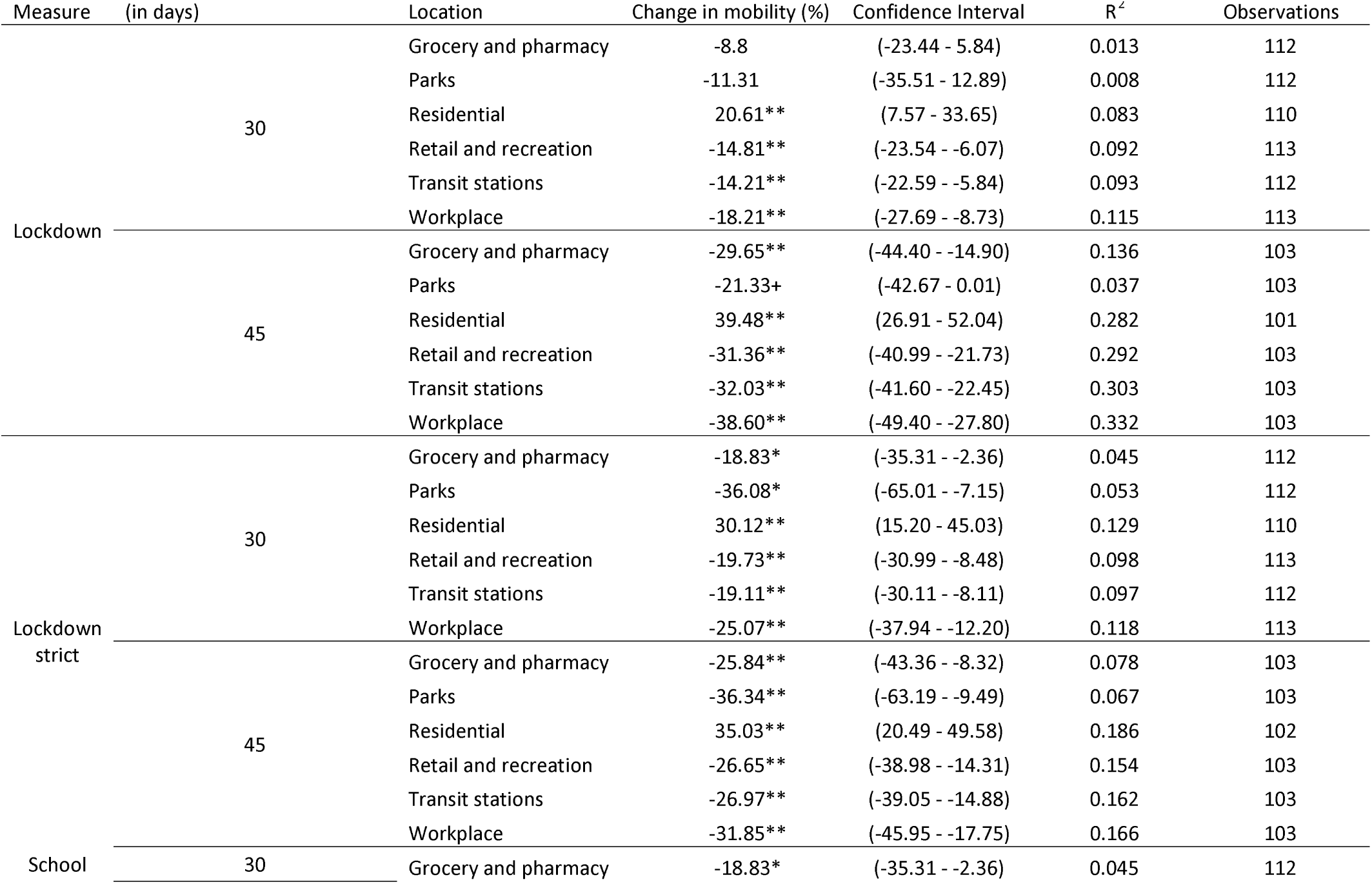
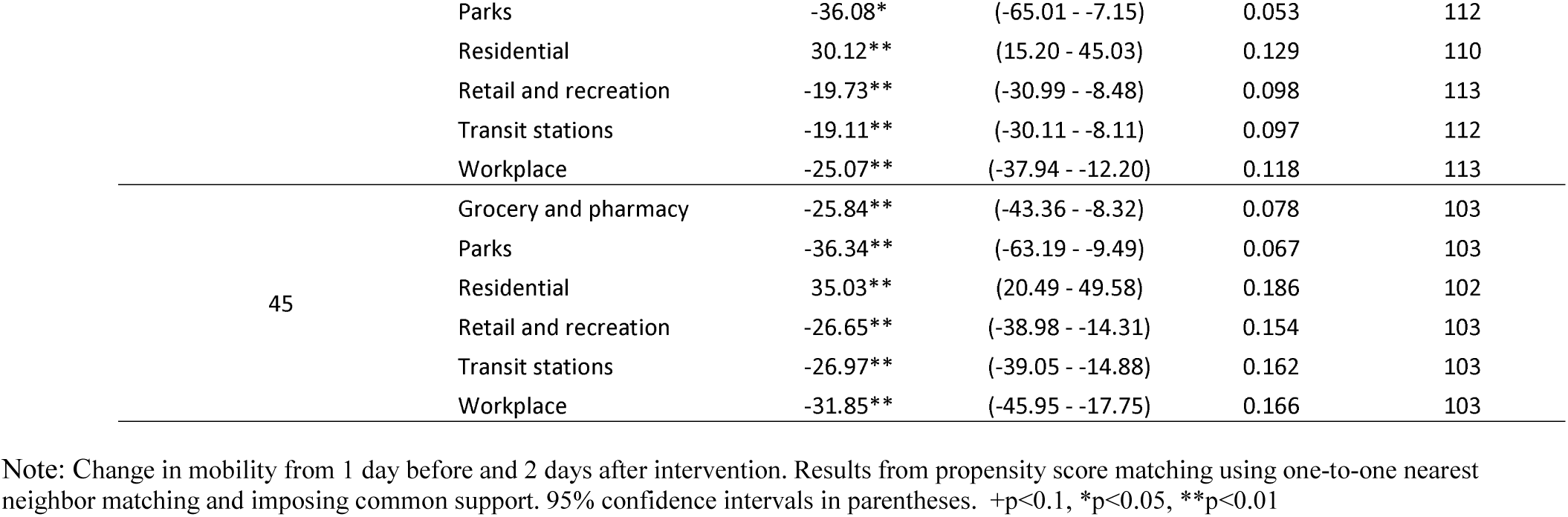
Propensity score matching results on change in mobility from the implementation of NPIs

| Measure | Latest day of implementation<br>(in days) | Location | Change in mobility (%) | Confidence Interval | R<sup>2</sup> | Observations |
| --- | --- | --- | --- | --- | --- | --- |
| Lockdown | 30 | Grocery and pharmacy | -8.8 | (-23.44 - 5.84) | 0.013 | 112 |
|  |  | Parks | -11.31 | (-35.51 - 12.89) | 0.008 | 112 |
|  |  | Residential | 20.61** | (7.57 - 33.65) | 0.083 | 110 |
|  |  | Retail and recreation | -14.81** | (-23.54 - -6.07) | 0.092 | 113 |
|  |  | Transit stations | -14.21** | (-22.59 - -5.84) | 0.093 | 112 |
|  |  | Workplace | -18.21** | (-27.69 - -8.73) | 0.115 | 113 |
|  | 45 | Grocery and pharmacy | -29.65** | (-44.40 - -14.90) | 0.136 | 103 |
|  |  | Parks | -21.33+ | (-42.67 - 0.01) | 0.037 | 103 |
|  |  | Residential | 39.48** | (26.91 - 52.04) | 0.282 | 101 |
|  |  | Retail and recreation | -31.36** | (-40.99 - -21.73) | 0.292 | 103 |
|  |  | Transit stations | -32.03** | (-41.60 - -22.45) | 0.303 | 103 |
|  |  | Workplace | -38.60** | (-49.40 - -27.80) | 0.332 | 103 |
| Lockdown<br>strict | 30 | Grocery and pharmacy | -18.83* | (-35.31 - -2.36) | 0.045 | 112 |
|  |  | Parks | -36.08* | (-65.01 - -7.15) | 0.053 | 112 |
|  |  | Residential | 30.12** | (15.20 - 45.03) | 0.129 | 110 |
|  |  | Retail and recreation | -19.73** | (-30.99 - -8.48) | 0.098 | 113 |
|  |  | Transit stations | -19.11** | (-30.11 - -8.11) | 0.097 | 112 |
|  |  | Workplace | -25.07** | (-37.94 - -12.20) | 0.118 | 113 |
|  | 45 | Grocery and pharmacy | -25.84** | (-43.36 - -8.32) | 0.078 | 103 |
|  |  | Parks | -36.34** | (-63.19 - -9.49) | 0.067 | 103 |
|  |  | Residential | 35.03** | (20.49 - 49.58) | 0.186 | 102 |
|  |  | Retail and recreation | -26.65** | (-38.98 - -14.31) | 0.154 | 103 |
|  |  | Transit stations | -26.97** | (-39.05 - -14.88) | 0.162 | 103 |
|  |  | Workplace | -31.85** | (-45.95 - -17.75) | 0.166 | 103 |
| School | 30 | Grocery and pharmacy | -18.83* | (-35.31 - -2.36) | 0.045 | 112 |

|  |  |  |  |  |  |
| --- | --- | --- | --- | --- | --- |
|  | Parks | -36.08* | (-65.01 - -7.15) | 0.053 | 112 |
|  | Residential | 30.12** | (15.20 - 45.03) | 0.129 | 110 |
|  | Retail and recreation | -19.73** | (-30.99 - -8.48) | 0.098 | 113 |
|  | Transit stations | -19.11** | (-30.11 - -8.11) | 0.097 | 112 |
|  | Workplace | -25.07** | (-37.94 - -12.20) | 0.118 | 113 |
| 45 | Grocery and pharmacy | -25.84** | (-43.36 - -8.32) | 0.078 | 103 |
|  | Parks | -36.34** | (-63.19 - -9.49) | 0.067 | 103 |
|  | Residential | 35.03** | (20.49 - 49.58) | 0.186 | 102 |
|  | Retail and recreation | -26.65** | (-38.98 - -14.31) | 0.154 | 103 |
|  | Transit stations | -26.97** | (-39.05 - -14.88) | 0.162 | 103 |
|  | Workplace | -31.85** | (-45.95 - -17.75) | 0.166 | 103 |

Note: Change in mobility from 1 day before and 2 days after intervention. Results from propensity score matching using one-to-one nearest neighbor matching and imposing common support. 95% confidence intervals in parentheses. +p<0.1, \*p<0.05, \*\*p<0.01

Tables A1 to A3 in the supplementary appendix show measures of balance and the ability of the matching model to reduce systematic differences between the treatment and control variables to make both groups comparable for analysis. Balancing tests are shown for all location categories because some countries were missing location data on some days, which may make matching results different across models for each location. For normal and strict lockdowns, the PSM procedure decreased differences between the treated and untreated groups after matching. There were substantial reductions in mean and median percentage difference between control and treated groups on matching variables. The p-value of the joint significance test was higher and insignificant in the matched sample and the pseudo R^2^ was substantially higher. For measuring the effect of school closure implemented before 30 days, there was also evidence that balance was achieved between the treated and control groups. However, when analyzing school closures implemented before 45 days, the joint significance test was still significant after the matching procedure, which may suggest that balance was not achieved in these models, and potential bias in our results.

## DISCUSSION

Our results show that there may be health capacity, socioeconomic, and epidemiological factors that determine which NPIs a country implements and the timing of interventions. Furthermore, we found that the implementation of lockdowns does reduce mobility, however, implementation of lockdowns backed by measures such as curfews or fines can be more effective in reducing mobility. Weak stay-at-home orders that that merely suggest working at home and only leaving when absolutely necessary relative to orders that strictly define when an individual can leave home, reduce mobility, but at lower rates, especially in earlier periods of an epidemic. Stricter lockdowns can also result in greater sustained reduced mobility over longer periods.

Countries with higher population density, higher income level, and later first case detection were more likely to implement NPIs. Population density is a risk factor for transmission, where more crowding and contact, can increase the rate of transmission. Delayed arrival of COVID-19 may give countries time to prepare for implementation and garner public support for interventions. Higher income countries may be able to absorb the costs of NPIs and provide social safety nets for their citizens. National lockdowns may not be feasible in poorer countries where support systems do not exist and large segments of the population are daily wage workers.

Countries with a higher number of cases per capita two weeks after first case detection were less likely to implement measures. While countries with greater transmission may have greater incentive to implement NPIs, we believe that initial growth in cases here may proxy for testing ability of the country and general health system capacity. Countries with greater testing capacity, may have prioritized early case detection, contact tracing, and isolation and quarantine measures, and relied less on NPIs that have economic and social costs.^56^

For countries that implemented lockdowns, those with greater health security preparedness and higher income had a longer delay to implementation. The goal of NPIs is to reduce peak prevalence to ensure hospitals have the necessary equipment and health worker availability to handle patient load. Therefore, a country with greater health systems capacity may delay implementation of NPIs because they can respond to a higher number of peak infections without overwhelming the health system. Larger countries were found to have longer delays in implementation which may be due to logistical challenges in covering a very large population under an NPI.

We found that democratic countries were more likely to implement lockdowns. Existing legal frameworks and political systems can determine a country’s ability to implement measures nationally in a timely manner, including the ability to declare a national emergency, allocate resources towards diagnostics, prevention and treatment, or issue travel restrictions.^48,49^ Although autocratic regimes can generally implement measures quickly, in a crisis situation a state of emergency can centralize powers within the federal government.^50^ Furthermore, democratic governments are more accountable to their constituents, and therefore may take more proactive measures to protect their health and may already have better health infrastructure in place to meet this goal.^51^

Our results confirm that country-specific response and adherence to international guidance on NPIs depends on political and economic factors, and public health systems capacity.^52^ There should be increased funding for poorer countries during a pandemic to combat the economic costs of NPIs and encourage implementation. Incentives should be created so that countries with greater resources and health systems capacities do not significantly delay the implementation of NPIs in order to curb transmission early. Pandemic preparedness investments, related to early detection systems, laboratory diagnostic capacity, surveillance, and general health systems capacity, should be made globally to increase capacity to combat future disease outbreaks.^53^ These should meet international benchmarks, which should be continuously reviewed and revised. During a pandemic information sharing and research should be prioritized and incentivized to help limit transmission and the health toll, especially in countries where the disease outbreak initially spreads as these countries lack vital knowledge and data on how to best contain the outbreak in the most cost-effective manner.

We also found that strict lockdowns were more effective in reducing mobility relative to normal lockdowns immediately after lockdown implementation, especially when they were implemented earlier (within 30 days of first case detection in our analysis). This suggests that if a country wants strict and timely adherence to a lockdown measure during a pandemic, it should include fines, curfews, or additional enforcement mechanisms to increase adherence, or clearly delineate who may leave their residence and under what specific circumstances, rather than make mere recommendations, as compliance for the latter may be lower. When we included countries which implemented lockdowns within 45 days, the changes in mobility between strict lockdown and normal lockdown decreased, with possibly greater reductions with a normal lockdown. This may be because greater perceived risk susceptibility^54-59^ and belief in effectiveness of actions^57-59^ has been shown to increase preventive behavior, and both of these may increase with time as people are exposed to more information about the pandemic. Also, adherence to social distancing policies tends to increase as time of epidemic increases.^58,59^ However, when we considered a longer post-intervention period, there were even greater reductions in mobility outside of the home under the strict lockdown. This suggests that people may initially voluntarily heed government recommendations, but become complacent with time, and stricter lockdowns may be necessary if greater reductions in transmission are desired. The economic and social costs of stricter lockdown measures should be weighed against the decreases in disease transmission.

The effect of school closures on COVID-19 transmission and mortality has been extensively discussed in the literature.^44-47^ There is a concern that school closures may have unintended consequences. Children outside of school, combined with lack of daycare options may require caregivers outside of the home such as relatives, to travel to a child’s home to care for them. Conversely, parents or guardians may not go to work or work from home to care for children. Some of these guardians may also be healthcare workers, decreasing the critical supply of healthcare workers during a pandemic.^47^ Furthermore, students may continue to engage in social and physical contact outside of schools even after closures. Our results on mobility changes after national school closures showed no significant changes in physical mobility and this remains an area for further study.

To induce compliance to social distancing guidelines and decrease anxiety and stress in the public, good risk communication policies are essential.^11,60^ The risk communication strategy should be targeted to specific audiences considering the lens through which they process information, including considering the audiences’ political affilations^61,62^ and education levels^55,57,63^. Risk communication should be delivered on various platforms and should be consistent and frequent.^11^ The messaging should focus on how individual measures reduce spread and why they are important, backed by science and facts.^11^ A long-term strategy for the implementation of NPIs should consider the effect of communication on adherence — a survey of 894 residents in Italy found that surprise extensions of self-isolation guidelines resulted in decreased willingness to comply, specifically among residents who had higher compliance to previous quarantines.^64^

There are a number of important limitations to our study. First, there may be data censoring issues to consider. We only focused on countries that had either implemented policies within 30 or 45 days, or had at least 30 or 45 days pass since their first case as of May 1^st^, the last day we collected NPI data. For the latest day of data available on NPI implementation only five countries which included four island nations and one conflict area had not passed 30 days since first case detection. The country with the latest first case detection of these countries had 24 days pass since their first case. When we considered countries that had that reached their 45^th^ day since first case exposure, 42 countries were excluded, 22 of which had 40 to 44 days pass since their first case was detected, the majority of them being small island nations, followed by poorer African nations, and conflict areas. For many of these countries there was a lack of control variable data. Furthermore, our results showed that countries that had a later first case detection rate were more likely to implement NPIs. It is important to acknowledge the absence of these countries in our analysis, particularly for outcome variables where we considered policy implementation within 45 days. However, given the lack of control variable data on these countries and the unique country characteristics that may make these countries outliers in their response and the effects of the pandemic on them, we did not consider models for censored data.

There also may be data issue quality issues. Lack of testing capabilities and pandemic preparedness may have created a lag in detection of cases and caused a measurement error in our data. Furthermore, in some countries, stigmatization or overcrowding of health facilities may make it difficult to track disease arrival and spread as individuals fail to report health symptoms to authorities.^20^ Second, the google mobility reports are not perfect measures of mobility. Although smart phone use has significantly increased in past years, poorer countries or older populations may be less likely to use smartphones, resulting in measurement error in mobility. However, these may be the best measures of mobility we currently have. Third, we have excluded countries that implemented sub-national NPIs in our analysis due to data collection challenges. It can be argued that for a pandemic such as COVID-19, a patchwork response is not adequate to prevent transmission. However, we recognize that logistical, social, and political considerations may not allow for national NPI implementation. Future analysis should focus on the effects of sub-national policies on mobility.

Lastly, although we made efforts to distinguish between intensities of social distancing measures by looking at a *normal* lockdown relative to a *strict* lockdown, there are challenges in categorizing different types of NPIs due to the large variation in NPIs across countries. Our analysis also ignored complementary measures that were implemented such as restrictions on public and private transit or economic measures that compensated individuals which stayed home and did not work.

NPI implementation and its timing is based on an assessment of the potential health, economic, and social costs and benefits of different policy options. However, pandemics such as COVID-19 make the implementation of such measures global public goods. It is important to consider what constraints countries face and how resources can be better allocated to improve timely implementation of these measures. We provide evidence that these lockdowns, especially those backed by curfews and fines, or stricter stay-at-home orders can reduce physical mobility which may reduce transmission in the early stages of a pandemic. Future research should look specifically at the effects of these different measures on economic and social costs, and how different variations of these measures maximize reduction in transmission and minimize costs.

## Data Availability

Data are publicly available from published sources.

## SUPPLEMENTARY APPENDIX

**Table A1:** Balancing between matched and control groups from lockdown model

| Data | Maximum<br>implementation delay<br>(days) | Location | Pseudo R<sup>2</sup> | P>Chi2 | Mean bias (%) | Median bias (%) |
| --- | --- | --- | --- | --- | --- | --- |
| Matched | 30 | Grocery and<br>pharmacy | 0.11 | 0.58 | 14.99 | 12.09 |
| Unmatched |  |  | 0.28 | 0.00 | 24.80 | 23.27 |
| Matched |  | Parks | 0.11 | 0.58 | 14.99 | 12.09 |
| Unmatched |  |  | 0.28 | 0.00 | 24.80 | 23.27 |
| Matched |  | Residential | 0.43 | 0.00 | 22.74 | 21.27 |
| Unmatched |  |  | 0.28 | 0.00 | 26.45 | 25.90 |
| Matched |  | Retail and<br>recreation | 0.11 | 0.68 | 15.01 | 13.97 |
| Unmatched |  |  | 0.28 | 0.00 | 24.60 | 23.00 |
| Matched |  | Transit stations | 0.11 | 0.58 | 14.99 | 12.09 |
| Unmatched |  |  | 0.28 | 0.00 | 24.80 | 23.27 |
| Matched |  | Workplace | 0.11 | 0.68 | 15.01 | 13.97 |
| Unmatched |  |  | 0.28 | 0.00 | 24.60 | 23.00 |
| Matched | 45 | Grocery and<br>pharmacy | 0.10 | 0.57 | 11.33 | 9.99 |
| Unmatched |  |  | 0.27 | 0.00 | 28.03 | 28.08 |
| Matched |  | Parks | 0.10 | 0.57 | 11.33 | 9.99 |
| Unmatched |  |  | 0.27 | 0.00 | 28.03 | 28.08 |
| Matched |  | Residential | 0.13 | 0.31 | 13.83 | 13.63 |
| Unmatched |  |  | 0.27 | 0.00 | 29.40 | 32.87 |
| Matched |  | Retail and<br>recreation | 0.10 | 0.57 | 11.33 | 9.99 |
| Unmatched |  |  | 0.27 | 0.00 | 28.03 | 28.08 |
| Matched |  | Transit stations | 0.10 | 0.57 | 11.33 | 9.99 |
| Unmatched |  |  | 0.27 | 0.00 | 28.03 | 28.08 |
| Matched |  | Workplace | 0.10 | 0.57 | 11.33 | 9.99 |
| Unmatched |  |  | 0.27 | 0.00 | 28.03 | 28.08 |

Note: Results from propensity score matching using one-to-one nearest neighbor matching and imposing common support.

**Table A2:**
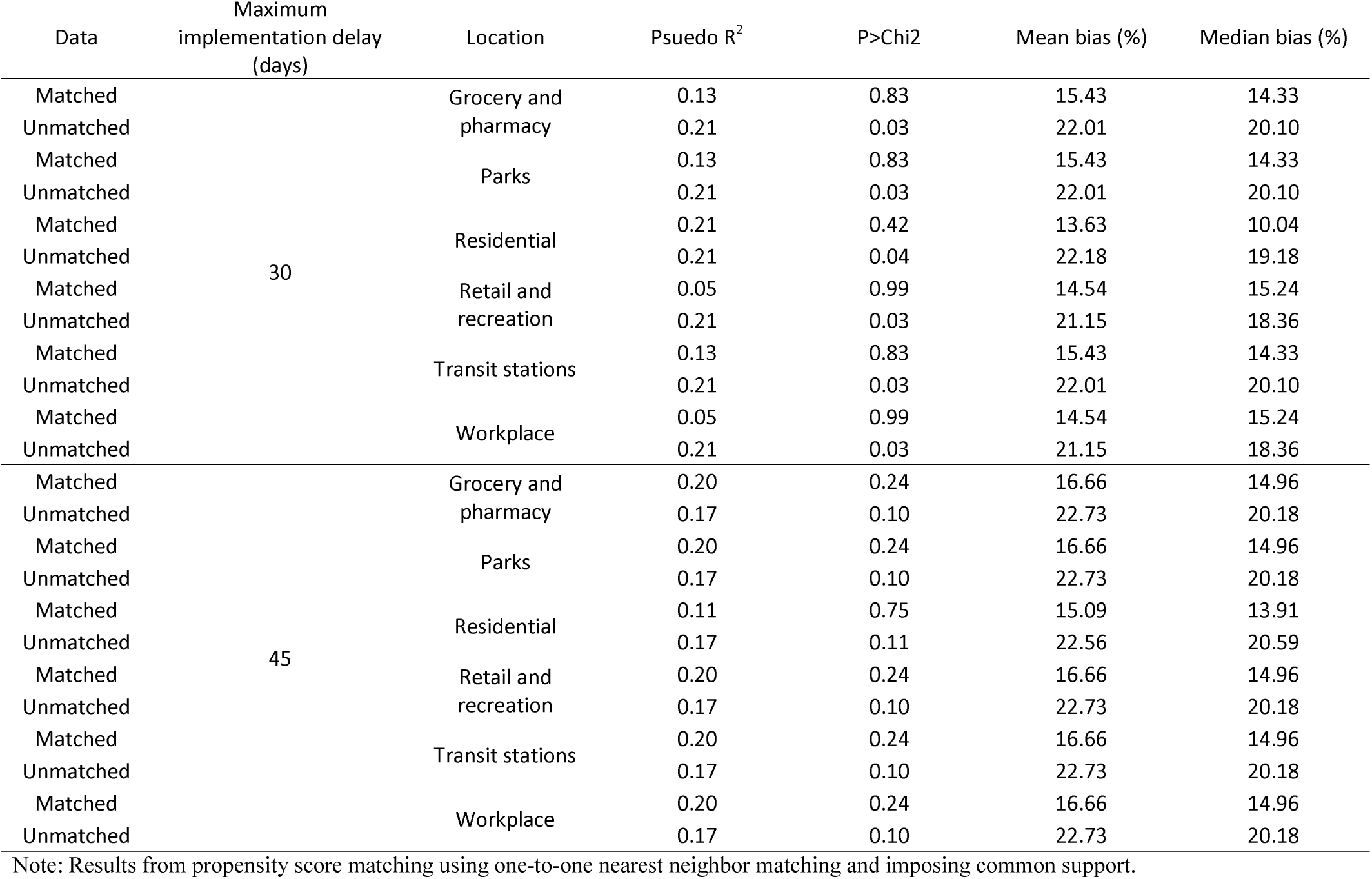
Balancing between matched and control groups from strict lockdown model

| Data | Maximum<br>implementation delay<br>(days) | Location | Pseudo R<sup>2</sup> | P>Chi2 | Mean bias (%) | Median bias (%) |
| --- | --- | --- | --- | --- | --- | --- |
| Matched | 30 | Grocery and<br>pharmacy | 0.13 | 0.83 | 15.43 | 14.33 |
| Unmatched |  |  | 0.21 | 0.03 | 22.01 | 20.10 |
| Matched |  | Parks | 0.13 | 0.83 | 15.43 | 14.33 |
| Unmatched |  |  | 0.21 | 0.03 | 22.01 | 20.10 |
| Matched |  | Residential | 0.21 | 0.42 | 13.63 | 10.04 |
| Unmatched |  |  | 0.21 | 0.04 | 22.18 | 19.18 |
| Matched |  | Retail and<br>recreation | 0.05 | 0.99 | 14.54 | 15.24 |
| Unmatched |  |  | 0.21 | 0.03 | 21.15 | 18.36 |
| Matched |  | Transit stations | 0.13 | 0.83 | 15.43 | 14.33 |
| Unmatched |  |  | 0.21 | 0.03 | 22.01 | 20.10 |
| Matched |  | Workplace | 0.05 | 0.99 | 14.54 | 15.24 |
| Unmatched |  |  | 0.21 | 0.03 | 21.15 | 18.36 |
| Matched | 45 | Grocery and<br>pharmacy | 0.20 | 0.24 | 16.66 | 14.96 |
| Unmatched |  |  | 0.17 | 0.10 | 22.73 | 20.18 |
| Matched |  | Parks | 0.20 | 0.24 | 16.66 | 14.96 |
| Unmatched |  |  | 0.17 | 0.10 | 22.73 | 20.18 |
| Matched |  | Residential | 0.11 | 0.75 | 15.09 | 13.91 |
| Unmatched |  |  | 0.17 | 0.11 | 22.56 | 20.59 |
| Matched |  | Retail and<br>recreation | 0.20 | 0.24 | 16.66 | 14.96 |
| Unmatched |  |  | 0.17 | 0.10 | 22.73 | 20.18 |
| Matched |  | Transit stations | 0.20 | 0.24 | 16.66 | 14.96 |
| Unmatched |  |  | 0.17 | 0.10 | 22.73 | 20.18 |
| Matched |  | Workplace | 0.20 | 0.24 | 16.66 | 14.96 |
| Unmatched |  |  | 0.17 | 0.10 | 22.73 | 20.18 |

Note: Results from propensity score matching using one-to-one nearest neighbor matching and imposing common support.

**Table A3:** Balancing between matched and control groups from school closure model

| Data | Maximum<br>implementation delay<br>(days) | Location | Pseudo R<sup>2</sup> | P>Chi2 | Mean bias (%) | Median bias (%) |
| --- | --- | --- | --- | --- | --- | --- |
| Matched | 30 | Grocery and<br>pharmacy | 0.14 | 0.23 | 14.91 | 13.65 |
| Unmatched |  |  | 0.57 | 0.00 | 44.74 | 37.78 |
| Matched |  | Parks | 0.14 | 0.23 | 14.91 | 13.65 |
| Unmatched |  |  | 0.57 | 0.00 | 44.74 | 37.78 |
| Matched |  | Residential | 0.16 | 0.18 | 14.40 | 13.42 |
| Unmatched |  |  | 0.57 | 0.00 | 44.99 | 37.15 |
| Matched |  | Retail and<br>recreation | 0.10 | 0.34 | 15.01 | 13.48 |
| Unmatched |  |  | 0.48 | 0.00 | 43.66 | 41.72 |
| Matched |  | Transit stations | 0.14 | 0.23 | 14.91 | 13.65 |
| Unmatched |  |  | 0.57 | 0.00 | 44.74 | 37.78 |
| Matched |  | Workplace | 0.10 | 0.34 | 15.01 | 13.48 |
| Unmatched |  |  | 0.48 | 0.00 | 43.66 | 41.72 |
| Matched | 45 | Grocery and<br>pharmacy | 0.33 | 0.00 | 26.36 | 25.57 |
| Unmatched |  |  | 0.59 | 0.00 | 56.25 | 55.75 |
| Matched |  | Parks | 0.33 | 0.00 | 26.36 | 25.57 |
| Unmatched |  |  | 0.59 | 0.00 | 56.25 | 55.75 |
| Matched |  | Residential | 0.20 | 0.01 | 22.30 | 15.80 |
| Unmatched |  |  | 0.58 | 0.00 | 56.50 | 55.11 |
| Matched |  | Retail and<br>recreation | 0.33 | 0.00 | 26.36 | 25.57 |
| Unmatched |  |  | 0.59 | 0.00 | 56.25 | 55.75 |
| Matched |  | Transit stations | 0.33 | 0.00 | 26.36 | 25.57 |
| Unmatched |  |  | 0.59 | 0.00 | 56.25 | 55.75 |
| Matched |  | Workplace | 0.33 | 0.00 | 26.36 | 25.57 |
| Unmatched |  |  | 0.59 | 0.00 | 56.25 | 55.75 |

Note: Results from propensity score matching using one-to-one nearest neighbor matching and imposing common support.

**Table A4:** Propensity score matching results on change in mobility from the implementation of lockdown (alternate matching)

| Latest delay in implementation (in days) | Matching model | Location | Change in mobility (%) | Confidence Interval | R<sup>2</sup> | Observations |
| --- | --- | --- | --- | --- | --- | --- |
| 30 | Nearest three neighbors | Grocery and pharmacy | -8.8 | (-23.44 - 5.84) | 0.013 | 112 |
|  | Kernel |  | -8.8 | (-23.44 - 5.84) | 0.013 | 112 |
|  | Nearest three neighbors | Parks | -11.31 | (-35.51 - 12.89) | 0.008 | 112 |
|  | Kernel |  | -11.31 | (-35.51 - 12.89) | 0.008 | 112 |
|  | Nearest three neighbors | Residential | 20.61** | (7.57 - 33.65) | 0.083 | 110 |
|  | Kernel |  | 20.61** | (7.57 - 33.65) | 0.083 | 110 |
|  | Nearest three neighbors | Retail and recreation | -14.81** | (-23.54 - -6.07) | 0.092 | 113 |
|  | Kernel |  | -14.81** | (-23.54 - -6.07) | 0.092 | 113 |
|  | Nearest three neighbors | Transit stations | -14.21** | (-22.59 - -5.84) | 0.093 | 112 |
|  | Kernel |  | -14.21** | (-22.59 - -5.84) | 0.093 | 112 |
|  | Nearest three neighbors | Workplace | -18.21** | (-27.69 - -8.73) | 0.115 | 113 |
|  | Kernel |  | -18.21** | (-27.69 - -8.73) | 0.115 | 113 |
| 45 | Nearest three neighbors | Grocery and pharmacy | -29.65** | (-44.40 - -14.90) | 0.136 | 103 |
|  | Kernel |  | -29.65** | (-44.40 - -14.90) | 0.136 | 103 |
|  | Nearest three neighbors | Parks | -21.33+ | (-42.67 - 0.01) | 0.037 | 103 |
|  | Kernel |  | -21.33+ | (-42.67 - 0.01) | 0.037 | 103 |
|  | Nearest three neighbors | Residential | 39.48** | (26.91 - 52.04) | 0.282 | 101 |
|  | Kernel |  | 39.48** | (26.91 - 52.04) | 0.282 | 101 |
|  | Nearest three neighbors | Retail and recreation | -31.36** | (-40.99 - -21.73) | 0.292 | 103 |
|  | Kernel |  | -31.36** | (-40.99 - -21.73) | 0.292 | 103 |
|  | Nearest three neighbors | Transit stations | -32.03** | (-41.60 - -22.45) | 0.303 | 103 |
|  | Kernel |  | -32.03** | (-41.60 - -22.45) | 0.303 | 103 |
|  | Nearest three neighbors | Workplace | -38.60** | (-49.40 - -27.80) | 0.332 | 103 |
|  | Kernel |  | -38.60** | (-49.40 - -27.80) | 0.332 | 103 |

Note: Outcome is change in mobility from 1 day before and 2 days after intervention. Common support imposed. 95% confidence intervals in parentheses. +p<0.1, \*p<0.05, \*\*p<0.01.

**Table A5:**
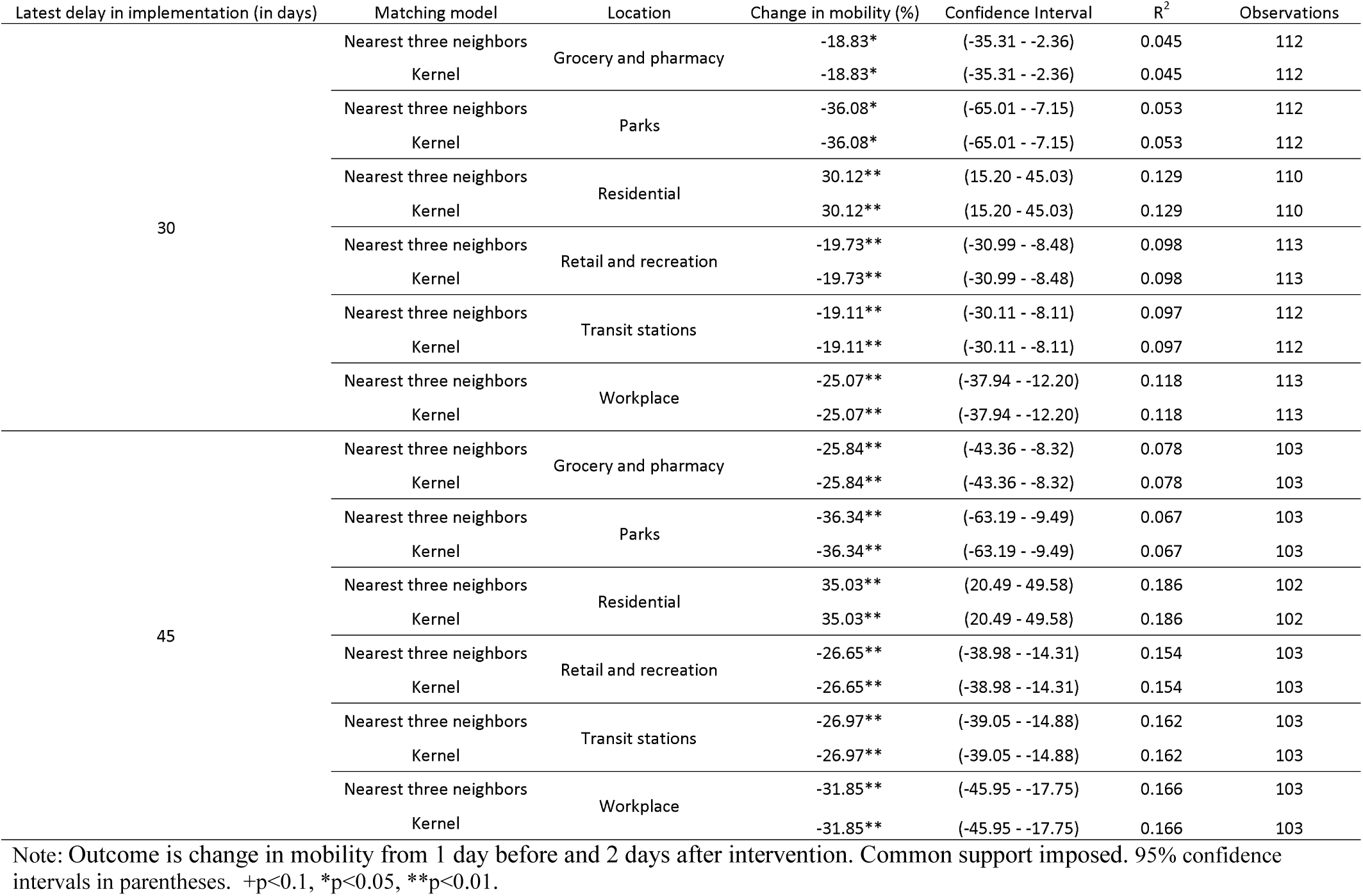
Propensity score matching results on change in mobility from the implementation of strict lockdown (alternate matching)

| Latest delay in implementation (in days) | Matching model | Location | Change in mobility (%) | Confidence Interval | R<sup>2</sup> | Observations |
| --- | --- | --- | --- | --- | --- | --- |
| 30 | Nearest three neighbors | Grocery and pharmacy | -18.83* | (-35.31 - -2.36) | 0.045 | 112 |
|  | Kernel |  | -18.83* | (-35.31 - -2.36) | 0.045 | 112 |
|  | Nearest three neighbors | Parks | -36.08* | (-65.01 - -7.15) | 0.053 | 112 |
|  | Kernel |  | -36.08* | (-65.01 - -7.15) | 0.053 | 112 |
|  | Nearest three neighbors | Residential | 30.12** | (15.20 - 45.03) | 0.129 | 110 |
|  | Kernel |  | 30.12** | (15.20 - 45.03) | 0.129 | 110 |
|  | Nearest three neighbors | Retail and recreation | -19.73** | (-30.99 - -8.48) | 0.098 | 113 |
|  | Kernel |  | -19.73** | (-30.99 - -8.48) | 0.098 | 113 |
|  | Nearest three neighbors | Transit stations | -19.11** | (-30.11 - -8.11) | 0.097 | 112 |
|  | Kernel |  | -19.11** | (-30.11 - -8.11) | 0.097 | 112 |
|  | Nearest three neighbors | Workplace | -25.07** | (-37.94 - -12.20) | 0.118 | 113 |
|  | Kernel |  | -25.07** | (-37.94 - -12.20) | 0.118 | 113 |
| 45 | Nearest three neighbors | Grocery and pharmacy | -25.84** | (-43.36 - -8.32) | 0.078 | 103 |
|  | Kernel |  | -25.84** | (-43.36 - -8.32) | 0.078 | 103 |
|  | Nearest three neighbors | Parks | -36.34** | (-63.19 - -9.49) | 0.067 | 103 |
|  | Kernel |  | -36.34** | (-63.19 - -9.49) | 0.067 | 103 |
|  | Nearest three neighbors | Residential | 35.03** | (20.49 - 49.58) | 0.186 | 102 |
|  | Kernel |  | 35.03** | (20.49 - 49.58) | 0.186 | 102 |
|  | Nearest three neighbors | Retail and recreation | -26.65** | (-38.98 - -14.31) | 0.154 | 103 |
|  | Kernel |  | -26.65** | (-38.98 - -14.31) | 0.154 | 103 |
|  | Nearest three neighbors | Transit stations | -26.97** | (-39.05 - -14.88) | 0.162 | 103 |
|  | Kernel |  | -26.97** | (-39.05 - -14.88) | 0.162 | 103 |
|  | Nearest three neighbors | Workplace | -31.85** | (-45.95 - -17.75) | 0.166 | 103 |
|  | Kernel |  | -31.85** | (-45.95 - -17.75) | 0.166 | 103 |

Note: Outcome is change in mobility from 1 day before and 2 days after intervention. Common support imposed. 95% confidence intervals in parentheses. +p<0.1, \*p<0.05, \*\*p<0.01.

**Table A6:** Propensity score matching results on change in mobility from the implementation of school closure (alternate matching)

| Latest delay in implementation (in days) | Matching model | Location | Change in mobility (%) | Confidence Interval | R<sup>2</sup> | Observations |
| --- | --- | --- | --- | --- | --- | --- |
| 30 | Nearest three neighbors | Grocery and pharmacy | -7.38 | (-21.98 - 7.21) | 0.011 | 96 |
|  | Kernel |  | -7.38 | (-21.98 - 7.21) | 0.011 | 96 |
|  | Nearest three neighbors | Parks | -10.58 | (-31.15 - 10.00) | 0.011 | 96 |
|  | Kernel |  | -10.58 | (-31.15 - 10.00) | 0.011 | 96 |
|  | Nearest three neighbors | Residential | 10.39 | (-3.41 - 24.19) | 0.024 | 94 |
|  | Kernel |  | 10.39 | (-3.41 - 24.19) | 0.024 | 94 |
|  | Nearest three neighbors | Retail and recreation | -15.25** | (-26.74 - -3.75) | 0.069 | 96 |
|  | Kernel |  | -15.25** | (-26.74 - -3.75) | 0.069 | 96 |
|  | Nearest three neighbors | Transit stations | -16.26** | (-28.18 - -4.35) | 0.073 | 96 |
|  | Kernel |  | -16.26** | (-28.18 - -4.35) | 0.073 | 96 |
|  | Nearest three neighbors | Workplace | -17.82** | (-30.92 - -4.72) | 0.072 | 96 |
|  | Kernel |  | -17.82** | (-30.92 - -4.72) | 0.072 | 96 |
| 45 | Nearest three neighbors | Grocery and pharmacy | -5.13 | (-20.33 - 10.08) | 0.005 | 90 |
|  | Kernel |  | -5.13 | (-20.33 - 10.08) | 0.005 | 90 |
|  | Nearest three neighbors | Parks | -7.91 | (-29.68 - 13.85) | 0.006 | 90 |
|  | Kernel |  | -7.91 | (-29.68 - 13.85) | 0.006 | 90 |
|  | Nearest three neighbors | Residential | 6.52 | (-7.92 - 20.95) | 0.009 | 88 |
|  | Kernel |  | 6.52 | (-7.92 - 20.95) | 0.009 | 88 |
|  | Nearest three neighbors | Retail and recreation | -13.45* | (-25.56 - -1.34) | 0.052 | 90 |
|  | Kernel |  | -13.45* | (-25.56 - -1.34) | 0.052 | 90 |
|  | Nearest three neighbors | Transit stations | -15.47* | (-27.98 - -2.96) | 0.064 | 90 |
|  | Kernel |  | -15.47* | (-27.98 - -2.96) | 0.064 | 90 |
|  | Nearest three neighbors | Workplace | -14.70* | (-28.38 - -1.02) | 0.049 | 90 |
|  | Kernel |  | -14.70* | (-28.38 - -1.02) | 0.049 | 90 |

Note: Outcome is change in mobility from 1 day before and 2 days after intervention. Common support imposed. 95% confidence intervals in parentheses. +p<0.1, \*p<0.05, \*\*p<0.01.

**Table A7:** Propensity score matching results on change in mobility from the implementation of NPIs (long follow-up)

| Measure | Latest day of implementation | Location | Change in mobility (%) | Confidence Interval | R<sup>2</sup> | Observations |
| --- | --- | --- | --- | --- | --- | --- |
| Lockdown | 30 | Grocery and pharmacy | -16.90** | (-27.80 - -6.01) | 0.079 | 112 |
|  |  | Parks | 4.42 | (-104.60 - 113.44) | 0 | 112 |
|  |  | Residential | 141.14** | (71.79 - 210.48) | 0.133 | 108 |
|  |  | Retail and recreation | -7.77** | (-12.14 - -3.41) | 0.101 | 113 |
|  |  | Transit stations | -7.23** | (-11.79 - -2.67) | 0.082 | 112 |
|  |  | Workplace | -14.23** | (-21.24 - -7.23) | 0.127 | 113 |
|  | 45 | Grocery and pharmacy | -39.46** | (-51.17 - -27.75) | 0.307 | 103 |
|  |  | Parks | 12.46 | (-88.57 - 113.49) | 0.001 | 103 |
|  |  | Residential | 198.65** | (133.05 - 264.24) | 0.269 | 100 |
|  |  | Retail and recreation | -15.84** | (-22.26 - -9.42) | 0.192 | 103 |
|  |  | Transit stations | -16.17** | (-23.22 - -9.11) | 0.17 | 103 |
|  |  | Workplace | -26.62** | (-35.70 - -17.54) | 0.251 | 103 |
| Lockdown strict | 30 | Grocery and pharmacy | -30.46** | (-47.74 - -13.18) | 0.1 | 112 |
|  |  | Parks | -124.14 | (-293.13 - 44.85) | 0.019 | 112 |
|  |  | Residential | 203.15** | (127.41 - 278.88) | 0.207 | 110 |
|  |  | Retail and recreation | -10.14** | (-16.20 - -4.07) | 0.09 | 113 |
|  |  | Transit stations | -9.62** | (-16.32 - -2.91) | 0.068 | 112 |
|  |  | Workplace | -20.44** | (-32.14 - -8.74) | 0.097 | 113 |
|  | 45 | Grocery and pharmacy | -41.69** | (-62.44 - -20.94) | 0.136 | 103 |
|  |  | Parks | -95.15 | (-252.07 - 61.78) | 0.014 | 103 |
|  |  | Residential | 219.28** | (149.96 - 288.60) | 0.283 | 102 |
|  |  | Retail and recreation | -15.53** | (-24.23 - -6.82) | 0.11 | 103 |
|  |  | Transit stations | -14.26** | (-23.17 - -5.34) | 0.091 | 103 |
|  |  | Workplace | -26.49** | (-40.68 - -12.31) | 0.12 | 103 |
| School | 30 | Grocery and pharmacy | -11.67 | (-31.80 - 8.46) | 0.014 | 94 |
|  |  | Parks | -16.39 | (-128.17 - 95.39) | 0.001 | 94 |
|  |  | Residential | -27.88 | (-101.96 - 46.21) | 0.006 | 91 |
|  |  | Retail and recreation | -5.67 | (-16.65 - 5.32) | 0.01 | 106 |
|  |  | Transit stations | -2.21 | (-13.60 - 9.18) | 0.002 | 94 |
|  |  | Workplace | -0.92 | (-14.38 - 12.53) | 0 | 106 |
|  | 45 | Grocery and pharmacy | -20.63 | (-49.72 - 8.47) | 0.02 | 97 |
|  |  | Parks | -27.53 | (-142.59 - 87.53) | 0.002 | 97 |
|  |  | Residential | -51.69 | (-127.36 - 23.98) | 0.02 | 94 |
|  |  | Retail and recreation | -4.99 | (-16.26 - 6.29) | 0.008 | 97 |
|  |  | Transit stations | -1.02 | (-13.33 - 11.29) | 0 | 97 |
|  |  | Workplace | -8.28 | (-23.85 - 7.29) | 0.012 | 97 |

Note: Outcome is change in mobility from 1 day before and 6 days after intervention. Results from propensity score matching using one-to-one nearest neighbor matching and imposing common support. 95% confidence intervals in parentheses. +p<0.1, \*p<0.05, \*\*p<0.01.

**Table A8:**
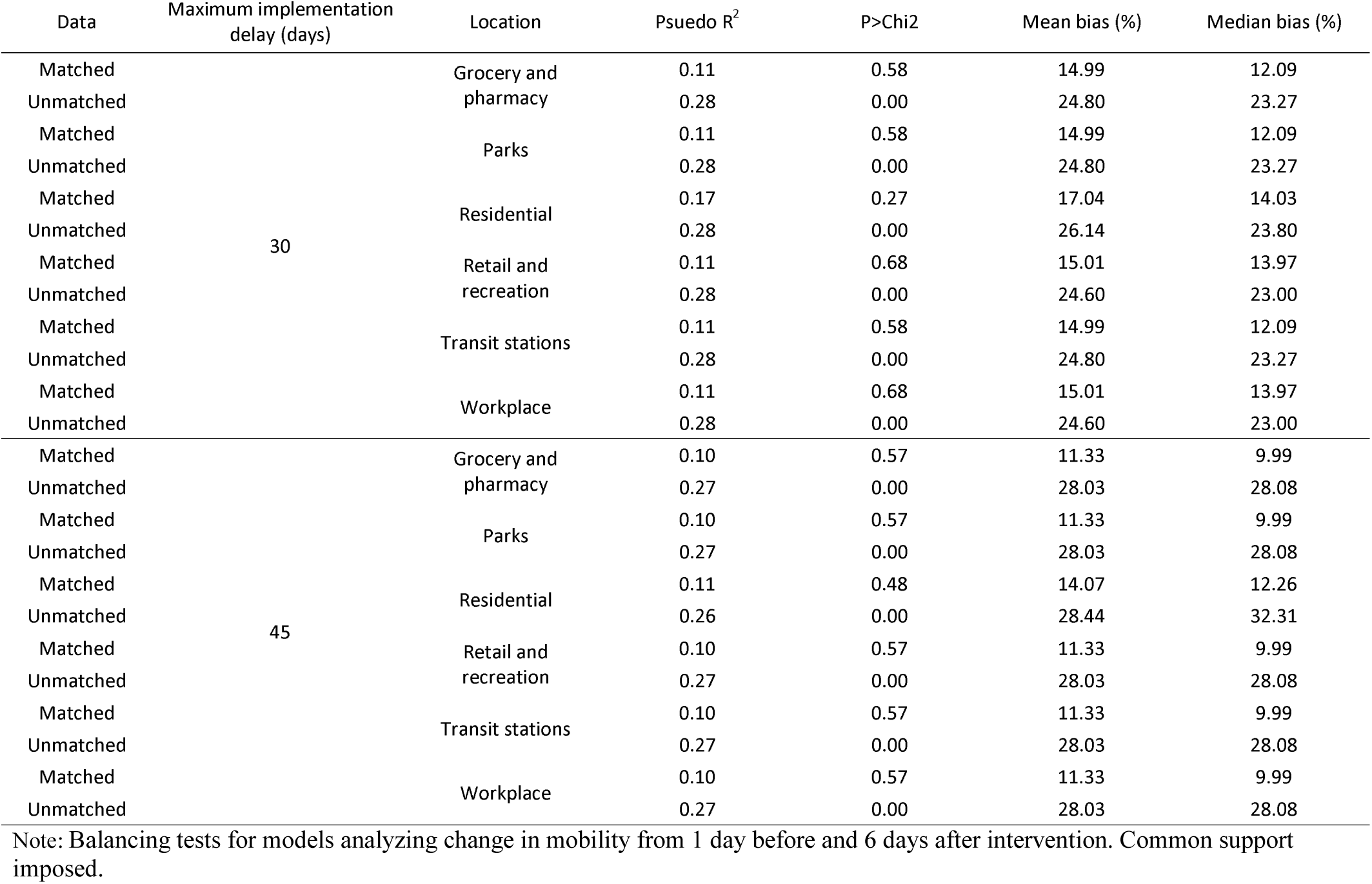
Balancing between matched and control groups from lockdown model (long follow-up)

| Data | Maximum implementation delay (days) | Location | Pseudo $R^2$ | P>Chi2 | Mean bias (%) | Median bias (%) |
| --- | --- | --- | --- | --- | --- | --- |
| Matched | 30 | Grocery and pharmacy | 0.11 | 0.58 | 14.99 | 12.09 |
| Unmatched |  |  | 0.28 | 0.00 | 24.80 | 23.27 |
| Matched |  | Parks | 0.11 | 0.58 | 14.99 | 12.09 |
| Unmatched |  |  | 0.28 | 0.00 | 24.80 | 23.27 |
| Matched |  | Residential | 0.17 | 0.27 | 17.04 | 14.03 |
| Unmatched |  |  | 0.28 | 0.00 | 26.14 | 23.80 |
| Matched |  | Retail and recreation | 0.11 | 0.68 | 15.01 | 13.97 |
| Unmatched |  |  | 0.28 | 0.00 | 24.60 | 23.00 |
| Matched |  | Transit stations | 0.11 | 0.58 | 14.99 | 12.09 |
| Unmatched |  |  | 0.28 | 0.00 | 24.80 | 23.27 |
| Matched |  | Workplace | 0.11 | 0.68 | 15.01 | 13.97 |
| Unmatched |  |  | 0.28 | 0.00 | 24.60 | 23.00 |
| Matched | 45 | Grocery and pharmacy | 0.10 | 0.57 | 11.33 | 9.99 |
| Unmatched |  |  | 0.27 | 0.00 | 28.03 | 28.08 |
| Matched |  | Parks | 0.10 | 0.57 | 11.33 | 9.99 |
| Unmatched |  |  | 0.27 | 0.00 | 28.03 | 28.08 |
| Matched |  | Residential | 0.11 | 0.48 | 14.07 | 12.26 |
| Unmatched |  |  | 0.26 | 0.00 | 28.44 | 32.31 |
| Matched |  | Retail and recreation | 0.10 | 0.57 | 11.33 | 9.99 |
| Unmatched |  |  | 0.27 | 0.00 | 28.03 | 28.08 |
| Matched |  | Transit stations | 0.10 | 0.57 | 11.33 | 9.99 |
| Unmatched |  |  | 0.27 | 0.00 | 28.03 | 28.08 |
| Matched |  | Workplace | 0.10 | 0.57 | 11.33 | 9.99 |
| Unmatched |  |  | 0.27 | 0.00 | 28.03 | 28.08 |

Note: Balancing tests for models analyzing change in mobility from 1 day before and 6 days after intervention. Common support imposed.

**Table A9:** Balancing between matched and control groups from strict lockdown model (long follow-up)

| Data | Maximum implementation delay (days) | Location | Pseudo $R^2$ | P>Chi2 | Mean bias (%) | Median bias (%) |
| --- | --- | --- | --- | --- | --- | --- |
| Matched | 30 | Grocery and pharmacy | 0.13 | 0.83 | 15.43 | 14.33 |
| Unmatched |  |  | 0.21 | 0.03 | 22.01 | 20.10 |
| Matched |  | Parks | 0.13 | 0.83 | 15.43 | 14.33 |
| Unmatched |  |  | 0.21 | 0.03 | 22.01 | 20.10 |
| Matched |  | Residential | 0.21 | 0.42 | 13.63 | 10.04 |
| Unmatched |  |  | 0.21 | 0.04 | 22.18 | 19.18 |
| Matched |  | Retail and recreation | 0.05 | 0.99 | 14.54 | 15.24 |
| Unmatched |  |  | 0.21 | 0.03 | 21.15 | 18.36 |
| Matched |  | Transit stations | 0.13 | 0.83 | 15.43 | 14.33 |
| Unmatched |  |  | 0.21 | 0.03 | 22.01 | 20.10 |
| Matched |  | Workplace | 0.05 | 0.99 | 14.54 | 15.24 |
| Unmatched |  |  | 0.21 | 0.03 | 21.15 | 18.36 |
| Matched | 45 | Grocery and pharmacy | 0.20 | 0.24 | 16.66 | 14.96 |
| Unmatched |  |  | 0.17 | 0.10 | 22.73 | 20.18 |
| Matched |  | Parks | 0.20 | 0.24 | 16.66 | 14.96 |
| Unmatched |  |  | 0.17 | 0.10 | 22.73 | 20.18 |
| Matched |  | Residential | 0.11 | 0.75 | 15.09 | 13.91 |
| Unmatched |  |  | 0.17 | 0.11 | 22.56 | 20.59 |
| Matched |  | Retail and recreation | 0.20 | 0.24 | 16.66 | 14.96 |
| Unmatched |  |  | 0.17 | 0.10 | 22.73 | 20.18 |
| Matched |  | Transit stations | 0.20 | 0.24 | 16.66 | 14.96 |
| Unmatched |  |  | 0.17 | 0.10 | 22.73 | 20.18 |
| Matched |  | Workplace | 0.20 | 0.24 | 16.66 | 14.96 |
| Unmatched |  |  | 0.17 | 0.10 | 22.73 | 20.18 |

Note: Balancing tests for models analyzing change in mobility from 1 day before and 6 days after intervention. Common support imposed.

**Table A10:** Balancing between matched and control groups from school closure model (long follow-up)

| Data | Maximum implementation delay (days) | Location | Pseudo $R^2$ | P>Chi2 | Mean bias (%) | Median bias (%) |
| --- | --- | --- | --- | --- | --- | --- |
| Matched | 30 | Grocery and pharmacy | 0.14 | 0.23 | 14.91 | 13.65 |
| Unmatched |  |  | 0.57 | 0.00 | 44.74 | 37.78 |
| Matched |  | Parks | 0.14 | 0.23 | 14.91 | 13.65 |
| Unmatched |  |  | 0.57 | 0.00 | 44.74 | 37.78 |
| Matched |  | Residential | 0.22 | 0.04 | 16.05 | 12.93 |
| Unmatched |  |  | 0.57 | 0.00 | 45.12 | 38.27 |
| Matched |  | Retail and recreation | 0.10 | 0.34 | 15.01 | 13.48 |
| Unmatched |  |  | 0.48 | 0.00 | 43.66 | 41.72 |
| Matched |  | Transit stations | 0.14 | 0.23 | 14.91 | 13.65 |
| Unmatched |  |  | 0.57 | 0.00 | 44.74 | 37.78 |
| Matched |  | Workplace | 0.10 | 0.34 | 15.01 | 13.48 |
| Unmatched |  |  | 0.48 | 0.00 | 43.66 | 41.72 |
| Matched | 45 | Grocery and pharmacy | 0.33 | 0.00 | 26.36 | 25.57 |
| Unmatched |  |  | 0.59 | 0.00 | 56.25 | 55.75 |
| Matched |  | Parks | 0.33 | 0.00 | 26.36 | 25.57 |
| Unmatched |  |  | 0.59 | 0.00 | 56.25 | 55.75 |
| Matched |  | Residential | 0.19 | 0.04 | 19.95 | 16.81 |
| Unmatched |  |  | 0.58 | 0.00 | 56.57 | 56.49 |
| Matched |  | Retail and recreation | 0.33 | 0.00 | 26.36 | 25.57 |
| Unmatched |  |  | 0.59 | 0.00 | 56.25 | 55.75 |
| Matched |  | Transit stations | 0.33 | 0.00 | 26.36 | 25.57 |
| Unmatched |  |  | 0.59 | 0.00 | 56.25 | 55.75 |
| Matched |  | Workplace | 0.33 | 0.00 | 26.36 | 25.57 |
| Unmatched |  |  | 0.59 | 0.00 | 56.25 | 55.75 |

Note: Balancing tests for models analyzing change in mobility from 1 day before and 6 days after intervention. Common support imposed.

